# Machine learning models to detect opioid misuse in Emergency Department patients at triage

**DOI:** 10.1101/2025.07.18.25331782

**Authors:** Chirag Chhablani, Usman Shahid, Natalie Parde, Sami Muslmani, Huiyi Hu, Dillon Thorpe, Majid Afshar, Niranjan Karnik, Neeraj Chhabra

## Abstract

**Objective:** Emergency department (ED) encounters represent valuable opportunities to initiate evidence-based treatments for patients with opioid misuse, but few receive such care. Universal manual screening has been proposed to improve patient identification but is uncommon due to its time and resource-intensive nature. We sought to determine the feasibility of identifying patients with opioid misuse at the time of ED triage using machine learning (ML).

**Methods:** We conducted a retrospective cohort study of 1,123 ED encounters (September 2020 – March 2023) at a tertiary hospital. Encounters were enriched for opioid misuse, manually annotated, and chronologically split for training, validation, and testing. Candidate triage-time features included patient demographics, Emergency Severity Index, arrival time of day, chief complaint, comorbidities, and chronic medications. Model performance was evaluated using F1 score, area under the precision–recall curve (AUPRC), accuracy, recall, and AUROC. Post-hoc explainability analyses included SHapley Additive exPlanations (SHAP) and feature importance.

**Results:** All models performed comparably to opioid-related diagnosis codes placed at any time during the encounter. Random Forest (F1=0.75 [95%CI 0.70-0.83], AUPRC=0.88 [0.81-0.93], accuracy=0.79 [0.70-0.83]) and Gradient Boosting (F1=0.77 [0.71-0.82], AUPRC=0.89 [0.85-0.93], accuracy=0.81 [0.720.84]) had among the highest F1 score and AUPRC but confidence intervals overlapped with other methods. Explainability analyses highlighted prior drug-use diagnosis codes, triage acuity, and age as top predictors.

**Conclusion:** ML classifiers leveraging routinely collected triage data offer a feasible alternative to manual screening in flagging opioid misuse before physician evaluation, potentially enabling early harm-reduction interventions. Prospective multi-site validation, calibration, and bias assessments are warranted.

## Introduction

Despite recent progress, the opioid crisis remains a public health emergency in the United States with overdose the leading cause of death for multiple age groups.^1,2^ The emergency department (ED) is a common first contact point between patients with opioid misuse and the broader healthcare system. Opioid-related ED encounters have increased with approximately 2 million opioid-related ED encounters between 2022 and 2024.^3^ Patients with opioid misuse disproportionately access emergency medical services, making these encounters valuable opportunities for the initiation of treatment and harm reduction measures.^4,5^ Accurate identification of opioid misuse during ED encounters is essential for treatment, care coordination, and resource allocation.

Despite the existence of evidence-based therapies, only a minority of ED patients with opioid misuse are offered treatment due to a variety of factors including varied patient presentations, receptivity to treatment, stigma, time constraints of care teams, and difficulties in patient identification and priortization.^6–8^ Universal manual screening for ED patients has been advocated to improve detection of patient with opioid misuse but has not been adopted on a broad scale as results from experiments have been mixed.^9–11^ Universal manual screening of patients entering the ED is a resource-intensive process requiring staff training and clinical workflow development which may conflict with other time-sensitive tasks and other manual screening tools which have already been implemented. Automated identification of patients via machine learning (ML) may offer a solution that does not take staff and resources away from other critical tasks.^12^ Language model-based approaches have shown promising results in identifying opioid misuse from clinical notes.^13–16^ These natural language processing-based algorithms for identifying opioid misuse require note processing and detection following clinical note completion which may only occur after a patient has physically left the ED. This limits the ability of ED staff to address dangerous opioid use while the patient is present in the ED. There remains a need for models designed for use early during an ED encounter allowing for interventions and care coordination in real-time.

The objective of this study was to determine the ability of machine learning classifiers in detecting opioid misuse using data already available or collected at the time of ED patient triage. We hypothesize that ML classifiers can accurately detect patients with opioid misuse, suggesting a possible role for automated screening at patient arrival in the ED immediately following the triage process.

## Methods

### Study Design and Cohort Development

This is a retrospective cohort study of ED patients at a single tertiary care center from September 2020 to March 2023. A cohort of 1200 ED encounters and corresponding data were extracted from the clinical data warehouse of the primary study institution. The sample included patients aged 18 years or older at the time of encounter. The sample was enriched for opioid misuse using previously described methods involving sampling ED encounters with a positive urine opiate screen without coexisting opioid prescription or administration or an opioid-related International Classification of Diseases, Tenth Revision, Clinical Modification (ICD-10-CM) diagnosis code and matching 1:1 with encounters lacking these criteria.^13,17,18^ Enrichment was pursued to create a balanced dataset for efficient model training, and matching was performed on disposition status rather than age and sex to minimize bias from expanded clinical documentation for admitted patients. Due to the low sensitivity of ICD-10-CM codes for detecting opioid misuse in the ED setting, we used expert manual annotation to assign ground truth labels for model training and testing.^17^ Opioid misuse is defined as use of opioids in any way that was not directed by a doctor and represents a heterogenous pattern of drug use that is difficult to categorize solely from structured variables in the EHR.^19^ We therefore followed a blinded and structured manual annotation schema using a definition for opioid misuse consistent with those from the National Institute of Drug Abuse (NIDA) and the National Survey on Drug Use and Health (NSDUH).^20,21^ Annotators underwent training and evaluation of interrater reliability with an expert prior to independent annotation and following completion of every 100 cases. The study was exempted as non-human subjects research by the primary study institution, was not pre-registered, and conforms, where appropriate, to the Transparent Reporting of a multivariable prediction model for Individual Prognosis Or Diagnosis, Artificial Intelligence (TRIPOD + AI) guidelines (Supplementary material).^22^

### Data Preprocessing

The dataset used in this study included a set of features available at the time of ED triage or entered during the triage process (Table 1). These features included both patient and encounter-specific variables. Patient-specific variables included demographics, past medical history, and medication lists. Encounter-specific variables included triage acuity, arrival time of day, and chief complaint. Triage acuity was measured by the Emergency Severity Index (ESI) which is used by the majority of EDs in the United States. The ESI is measured as a 5-point scale categorizing patients based on severity of their conditions from level 1 (most urgent) to level 5 (least urgent).^23,24^

**Table 1.**
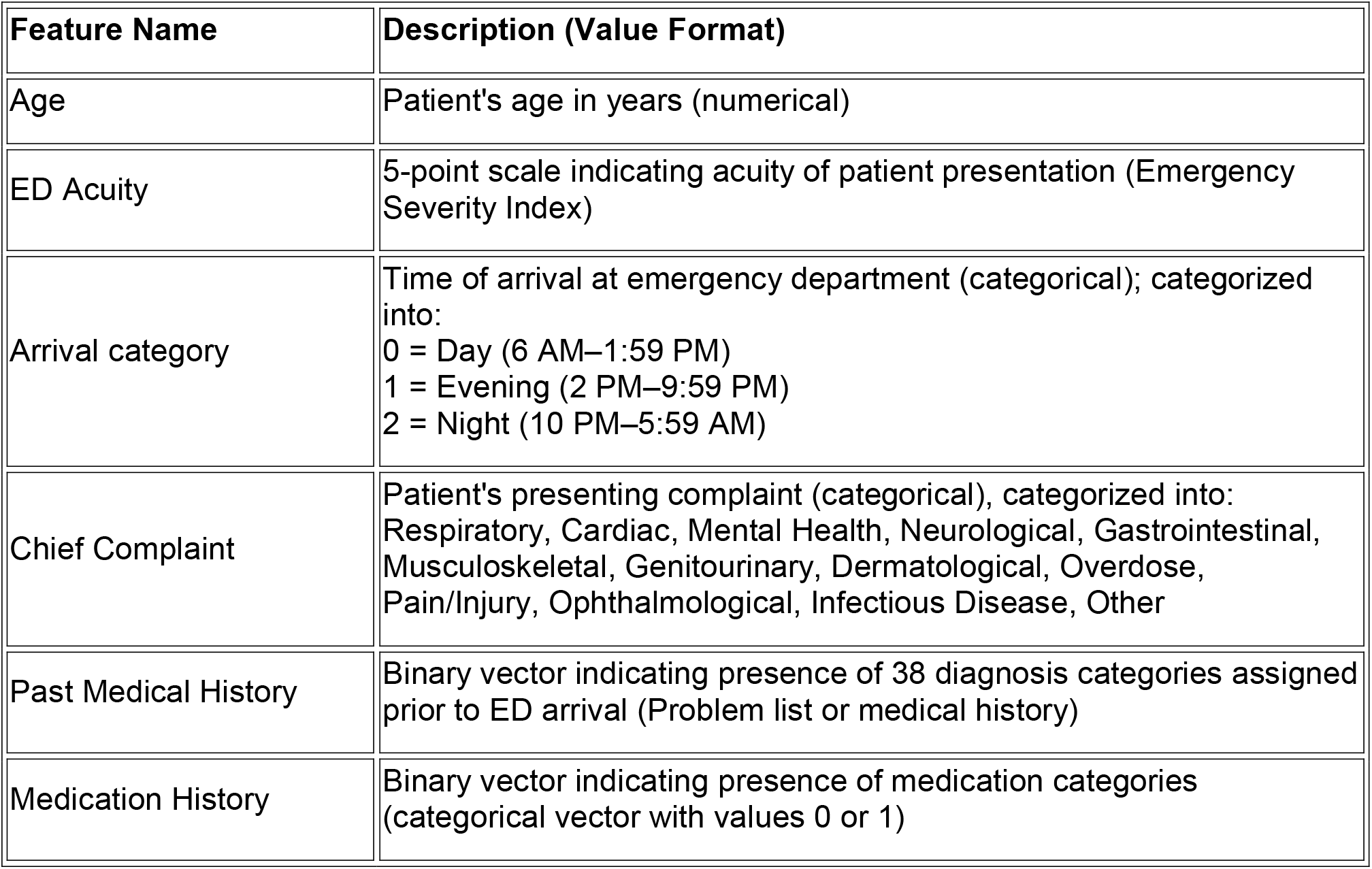
Candidate features for model inclusion.

| Feature Name | Description (Value Format) |
| --- | --- |
| Age | Patient's age in years (numerical) |
| ED Acuity | 5-point scale indicating acuity of patient presentation (Emergency Severity Index) |
| Arrival category | Time of arrival at emergency department (categorical); categorized into:<br>0 = Day (6 AM–1:59 PM)<br>1 = Evening (2 PM–9:59 PM)<br>2 = Night (10 PM–5:59 AM) |
| Chief Complaint | Patient's presenting complaint (categorical), categorized into: Respiratory, Cardiac, Mental Health, Neurological, Gastrointestinal, Musculoskeletal, Genitourinary, Dermatological, Overdose, Pain/Injury, Ophthalmological, Infectious Disease, Other |
| Past Medical History | Binary vector indicating presence of 38 diagnosis categories assigned prior to ED arrival (Problem list or medical history) |
| Medication History | Binary vector indicating presence of medication categories (categorical vector with values 0 or 1) |

All variables except for chief complaint represent structured data fields. Free-text chief complaints were preprocessed to enhance clinical interpretability and generalizability. We developed an expert-derived lexicon of keywords to map each complaint to one of several broad clinical categories: respiratory, cardiac, gastrointestinal, mental health, neurological, musculoskeletal, dermatological, drug use, pain/injury, genitourinary, ophthalmological, infectious disease, and other (see supplementary material). This keyword rule-based categorization allowed us to transform diverse textual complaints into a simple structured format that could be used as easily interpretable model inputs, while also offering a low-complexity approach requiring minimal computing resources that could be readily adapted to other settings. For medical history, we evaluated ICD-10-CM codes recorded prior to the ED arrival time, including those categorized as “Medical History” or as part of the “Problem List” with a designation of active. To reduce the dimensionality of medical history codes, we grouped codes into clinical categories using a standardized classification defined by the Elixhauser Comorbidity Index (ECI). The ECI maps comorbid diagnosis codes to 38 pre-existing condition categories and represents comorbidities that co-exist at the time of hospital encounter.^25–27^ Each category was converted into a binary feature, resulting in a structured representation of the patient’s diagnostic history leading up to the ED visit. Preprocessing categories for chief complaint and medical history are in the supplementary materials.

Each patient’s medication list was cleaned by removing dosage, frequency, and quantity. To reduce dimensionality, resulting medications were grouped into therapeutic classes defined by the United States Pharmacopeial Convention (USP) Drug Classification 2025 updated schema.^28^ Temporal patterns in ED arrivals were also encoded to capture potential circadian effects. The ED arrival time was discretized into three categories: day (6 AM to 1:59 PM), evening (2 PM to 9:59 PM), and night (10 PM to 5:59 AM). There was no missingness noted in variables apart from the ED acuity field, which contained 2 missing values (0.18% of the dataset). Given the minimal degree of missingness, we used replacement with median value. Results were robust to other methods of imputation (Supplement).

### Model development and explainability

The model training/testing cohort was developed using the sample of 1200 ED encounters, of which 77 were excluded due to enhanced privacy protections or lack of documentation in the clinical encounter. Encounters were sorted chronologically by date and divided into training, validation, and testing sets at a 70:15:15 ratio with the test set representing the most recent encounters to provide temporal validation and diminish the effects of selection bias due to COVID-related changes in healthcare utilization. We evaluated a diverse set of statistical and machine learning classifiers, including logistic regression with elastic net regularization, support vector machines (SVM), decision trees, K-nearest neighbors (KNN), naive bayes, bagging, random forest, gradient boosting (with different estimator settings), and AdaBoost. We also evaluated a feedforward neural network (FFNN). For our FFNN, we employed a fully connected feedforward neural network comprising three hidden layers with 128, 64, and 32 neurons respectively, each followed by ReLU activations and dropout for regularization. The final output layer produced a single logit for binary classification. Final model parameters are included in the supplementary materials. Models were evaluated along multiple metrics with the primary being Area Under the Precision Recall Curve (AUPRC), F1 score, precision (positive predictive value), recall (sensitivity), accuracy, and area under the receiver operator curve (AUROC). We used model classification threshold probability = 0.5 across all the classifiers. Confidence intervals were estimated using bootstrap resampling with replacement (n=1000 iterations). The 95% confidence intervals were defined as the 2.5th and 97.5th percentiles of the bootstrap distribution. Calibration curves were created for each model and included in the supplementary material. To provide context for the performance of our triage-only data models, we also calculated the performance of opioid-related ICD-10-CM diagnosis codes that were placed at any time during the encounter.^29^

We performed explainability analyses on the best performing classifier, selected by F1 score. Using SHAP (SHapley Additive exPlanations), we visualized the influence of individual features on model predictions in a beeswarm plot, with each point indicating one encounter. SHAP allows for both global and individual-level interpretability, showing the direction and magnitude of effect normalized to a 0-1 scale. We also examined global feature importance using random forest’s built-in Gini metric from scikit-learn, which ranks features by their contribution to overall classification.^30^ While Gini scores reflect general model behaviour, SHAP reveals how specific features drive individual predictions. Analyses were performed using Python (version 3.11). Given the use of protected health information, data cannot be shared but code will be shared with reasonable request to the study team.

## Results

There were 1123 ED encounters included for analysis with the most recent 169 encounters excluded from model training as the test set. Demographic information of the cohort is shown in Table 2. The opioid misuse cohort contained a greater proportion of male, White, Black, non-Hispanic, and Medicaid insured patients than the cohort without opioid misuse. Most of the triage-data only models performed comparably to the performance of ICD-10-CM codes placed at any time during the encounter (Table 3). Among machine learning models, ensemble methods such as Random Forest (Accuracy = 0.79, F1 Score = 0.75, AUROC = 0.87, AUPRC = 0.88) and Gradient Boosting (Accuracy = 0.81, F1 Score = 0.77, AUROC = 0.88, AUPRC = 0.89) achieved among the best overall performance of the classic ML methods but confidence intervals of most performance metrics overlapped with other model types including those for simpler models such as logistic regression.

**Table 2.** Demographic features.

| Feature | Opioid misuse<br>(n=570) | No opioid misuse<br>(n=568) |
| --- | --- | --- |
| Age (mean, SD) | 51.3 (14.0) | 46.5 (19.0) |
| Sex - Male | 379 (66.5%) | 207 (36.4%) |
| Sex - Female | 191 (33.5%) | 361 (63.6%) |
| Race - American Indian/Alaska Native | 1 (0.2%) | 3 (0.5%) |
| Race - Asian | 2 (0.4%) | 21 (3.5%) |
| Race - Black/African American | 347 (60.9%) | 308 (54.2%) |
| Race - White | 122 (21.4%) | 63 (11.1%) |
| Race - Hispanic | 88 (15.4%) | 169 (29.8%) |
| Race - Patient Declined | 10 (1.8%) | 4 (0.7%) |
| Ethnicity - Non-Hispanic or Latino | 465 (81.6%) | 388 (68.3%) |
| Ethnicity - Hispanic or Latino | 91 (16.0%) | 167 (29.4%) |
| Ethnicity - Patient Declined | 4 (0.7%) | 7 (1.2%) |
| Ethnicity - Unknown | 10 (1.8%) | 6 (1.1%) |
| Insurance Type - Private Insurance | 14 (2.5%) | 119 (21.0%) |
| Insurance Type - Medicaid | 395 (69.3%) | 281 (49.5%) |
| Insurance Type - Medicare | 126 (22.1%) | 119 (21.0%) |
| Insurance Type - Other | 10 (1.8%) | 8 (1.4%) |
| Insurance Type - No Information | 25 (4.4%) | 41 (7.2%) |

**Table 3.**
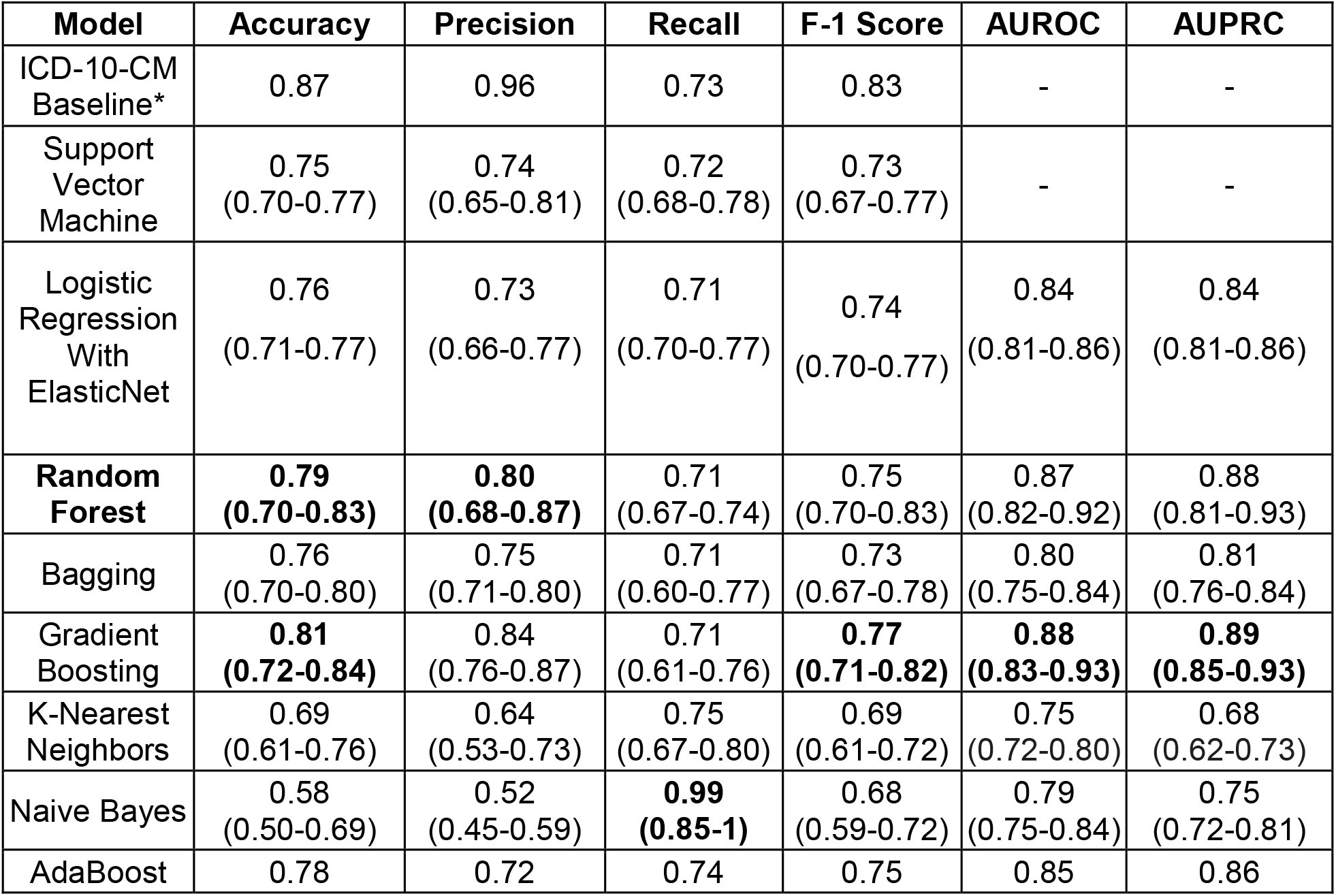
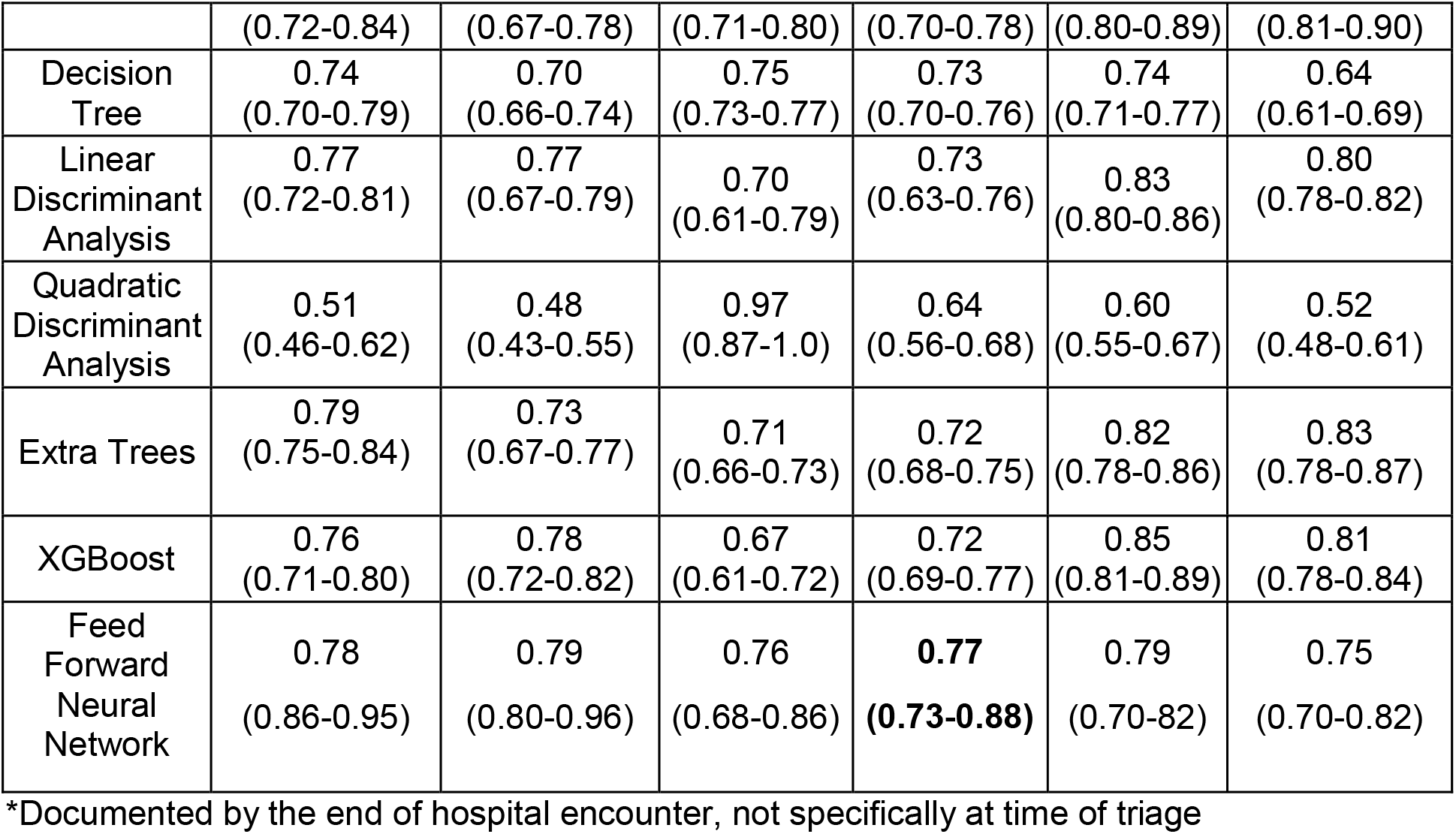
Model performance using triage-only data.

Model explainability analyses were performed on the random forest model (Figures 1 and 2). As the gradient boosting model had the highest AUPRC and FFNN had similar F1 score, we included the SHAP analyses for those models in the supplementary material. The presence of a prior drug use diagnosis code was top predictor across models. ED acuity and patient age were also among the top predictive features. Prior diagnosis codes for uncomplicated and complicated diabetes were negative predictors for opioid misuse while chronic pulmonary disease, alcohol abuse, and depression were positive predictive features. Non-daytime ED arrival times were also predictive of opioid misuse.

**Figure 1.**
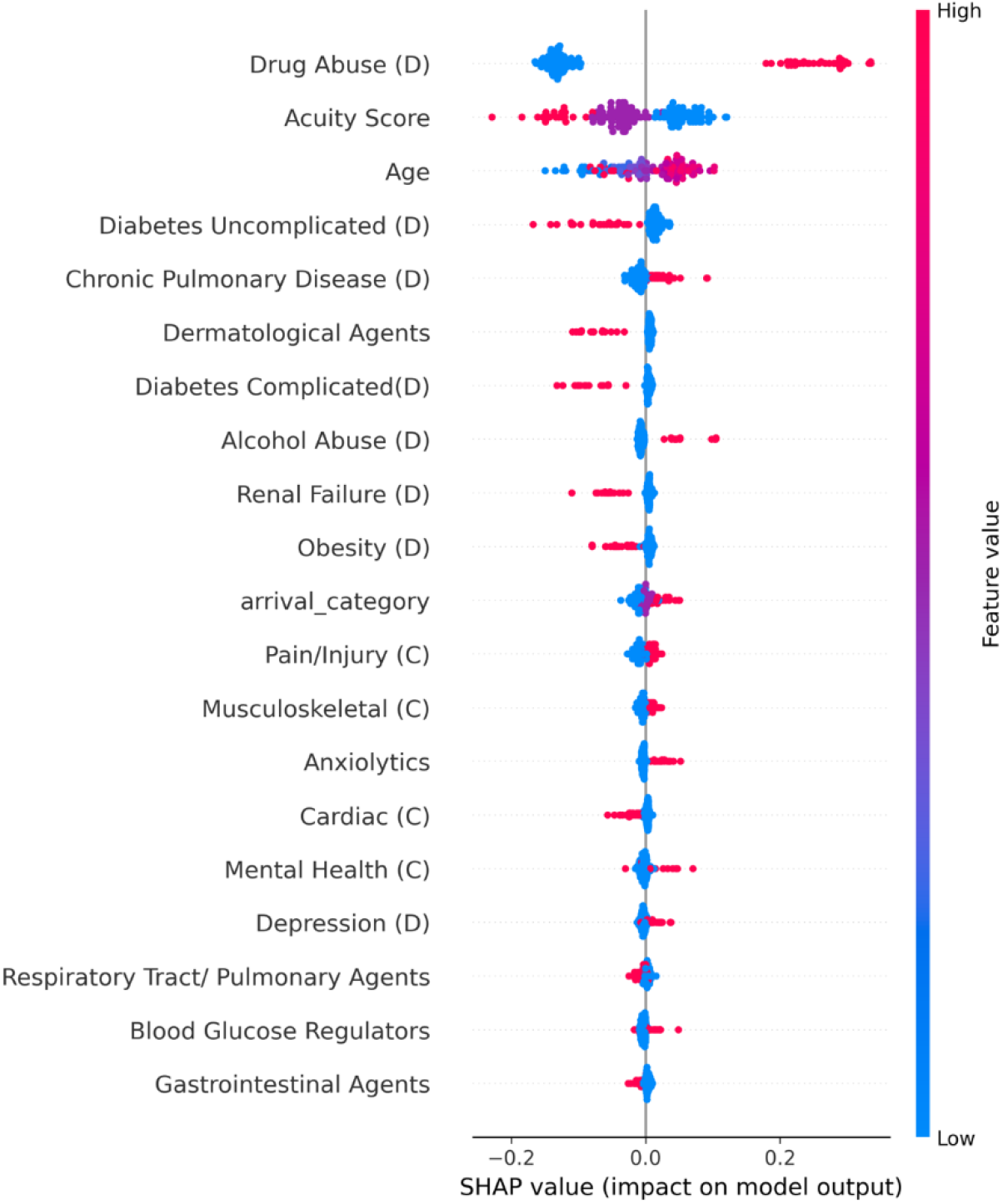
SHAP beeswarm plot for Random Forest Classifier

**Figure 2.**
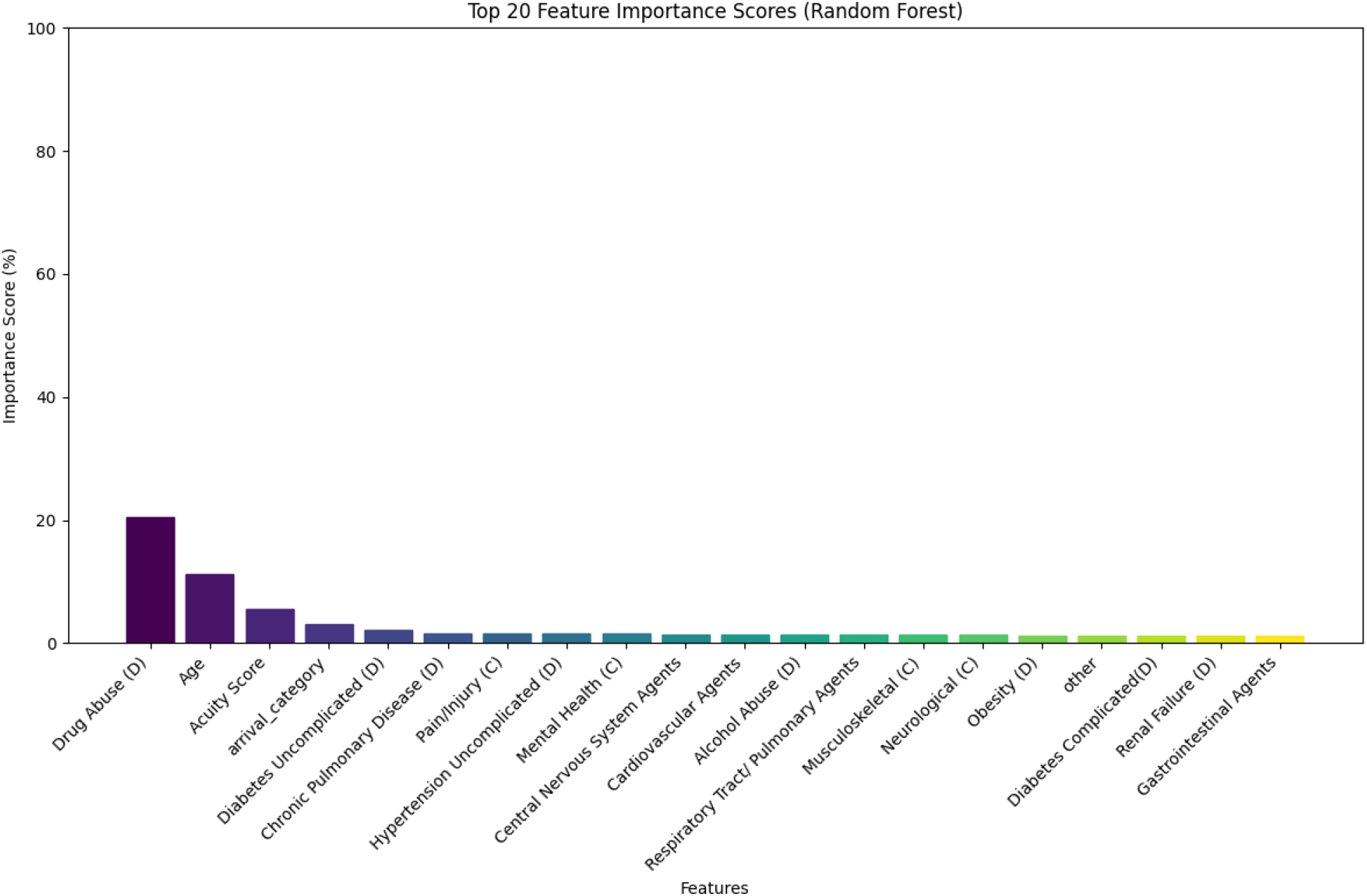
Global Feature Importance for Random Forest Classifier D=diagnosis category; C=chief complaint **Each point in the plot represents a single patient record. The x-axis shows SHAP value, reflecting the magnitude and direction of the feature’s impact on the model output: positive values indicate a strong predictor of opioid misuse classification, while negative values indicate a negative predictor. Points are color-coded according to the feature value, with red denoting opioid misuse and blue denoting no opioid misuse. Features are ordered by their average impact across the dataset. D=diagnosis category; C=chief complaint

### Limitations

All results should be interpreted within the context of their limitations. Data used in model training and testing were entered into the EHR for non-research purposes at a single institution and may not be universally representative. As ICD-10-CM codes were a portion of the criteria for cohort development, performance metrics for diagnosis codes were likely inflated. Given the limited sample size, evaluations for bias on demographic variables were uninformative. Models should undergo calibration and testing for bias and fairness in a larger cohort with true disease prevalence prior to testing in clinical trials.

## Discussion

These results demonstrate the ability of statistical and machine learning models to identify opioid misuse among ED patients using only data available within the EHR following patient triage. The sensitivity of models rivaled those of ICD-10-CM codes placed at any point during the encounter. Among the models tested, tree-based approaches like gradient boosting and random forest, achieved the best performance on the held-out test set but confidence intervals overlapped with other approaches. Feature importance evaluations revealed that pre-existing substance use-related diagnosis codes were the top predictors for opioid misuse. Given the limited sample of this pilot study, these results should be considered preliminary and confirmed in larger-scale evaluations on true prevalence for screening.

Top performing modeling included the decision tree-based approaches (random forest and gradient boosting models). These methods leverage the strength of multiple base learners, reducing variance and bias, and are particularly well-suited for structured datasets with heterogeneous feature importance. Simpler models like logistic regression and SVM yielded similar performance to the more computationally complex methods, including FFNN, indicating that relatively simpler approaches may have utility in this domain.

Explainability is critical for clinical machine learning models in healthcare to ensure transparency and trust. In our classifier, explainability analyses revealed that pre-existing diagnosis codes for opioid misuse were the top predictor, supporting the model’s face validity. Codes for alcohol use and depression were also predictive, reflecting known associations of opioid misuse with co-occurring substance use and mental health comorbidities.^31–33^ Chronic pulmonary disease also emerged as a key feature, possibly reflecting known links with insufflated opioids common in the location of the study.^34^ These findings highlight the value of both diagnostic history and comorbidities in substance use prediction. ED acuity had a mixed effect: high ESI (low acuity encounters) generally aligned with no misuse, but lower ESI did not consistently predict misuse.

Machine learning has the potential to automate clinical processes and reduce cognitive burden. The volume of EHR data can be overwhelming for clinicians, contributing to poor patient care and burnout.^35,36^ By leveraging routinely collected clinical data, machine learning models can surface relevant information in real time, streamlining workflows and reducing staff burden. Prior models using clinical text performed well but rely on data not available until later in the ED visit, limiting real-time utility.^13^ Our triage-based model provides a real-world proof of concept showing how EHR data can support timely disease detection in EDs.

To enhance generalizability and reduce the risk of overfitting in the context of our limited sample size, we incorporated preprocessing steps such as keyword searching and categorical encoding. While such preprocessing was done to improve model robustness, it introduced additional complexity that may hinder implementation. This tradeoff underscores the challenge of balancing performance with real-world feasibility. Future work should evaluate streamlined approaches that preserve accuracy with less operational burden.

Our findings suggest that predictive modeling using triage data can support early screening for opioid misuse which can be used to prompt diagnostic workups, harm reduction, or MOUD initiation. Such models could standardize and scale opioid misuse assessment as a routine part of ED care, potentially reducing variability and selection bias in practice. Prior to clinical deployment, model calibration and further evaluations of operational factors such as workup-to-detection ratios and numbers needed to evaluate are needed for safe integration into care. Future work should validate these models in larger cohorts and assess their impact on patient outcomes and clinical workflows.

We demonstrated the development and internal validation of high-performing machine learning models for opioid misuse screening. Future studies should evaluate the potential for triage-based machine learning models to improve care for patients with opioid misuse in the emergency department.

## Supporting information

TRIPOD

## Data Availability

All data produced in the present study are available upon reasonable request to the authors

