## Supplement for "Machine learning models to detect opioid misuse in Emergency Department patients at triage"

Supplementary material -

S. Table 1. Elixhauser Comorbidity Categories

| **#** | **Diagnostic Category** | **Abbreviation** | **ICD Codes** |
| --- | --- | --- | --- |
| 1 | Congestive Heart Failure | CHF | I099, I110, I130, I132, I255, I420, I425, I426, I427, I428, I429, I43, I50, P290 |
| 2 | Cardiac Arrhythmia | Arrhy | I441, I442, I443, I456, I459, I47, I48, I49, R000, R001, R008, T821, Z450, Z950 |
| 3 | Valvular Disease | VD | A520, I05, I06, I07, I08, I091, I098, I34, I35, I36, I37, I38, I39, Q230–Q233, Z952–Z954 |
| 4 | Pulmonary Circulation Disorders | PCD | I26, I27, I280, I288, I289 |
| 5 | Peripheral Vascular Disorders | PVD | I70, I71, I731, I738, I739, I771, I790, I792, K551, K558, K559, Z958, Z959 |
| 6 | Hypertension Uncomplicated | HPTN_NC | I10 |
| 7 | Hypertension Complicated | HPTN_C | I11, I12, I13, I15 |
| 8 | Paralysis | Para | G041, G114, G801, G802, G81, G82, G830–G834, G839 |
| 9 | Other Neurological Disorders | OthND | G10–G13, G20–G22, G254–G255, G312, G318–G319, G32, G35–G37, G40–G41, G931, G934, R470, R56 |
| 10 | Chronic Pulmonary Disease | COPD | I278, I279, J40–J47, J60–J67, J684, J701, J703 |
| 11 | Diabetes Uncomplicated | Diab_NC | E100–E101, E109–E111, E119, E120–E121, E129, E130–E131, E139, E140–E141, E149 |
| 12 | Diabetes Complicated | Diab_C | E102–E108, E112–E118, E122–E128, E132–E138, E142–E148 |
| 13 | Hypothyroidism | Hptothy | E00–E03, E890 |
| 14 | Renal Failure | RF | I120, I131, N18–N19, N250, Z490–Z492, Z940, Z992 |
| 15 | Liver Disease | LD | B18, I85, I864, I982, K70, K711, K713–K715, K717, K72–K74, K760–K769, Z944 |
| 16 | Peptic Ulcer Disease (excluding bleeding) | PUD_NB | K257, K259, K267, K269, K277, K279, K287, K289 |
| 17 | AIDS/HIV | HIV | B20, B21, B22, B24 |
| 18 | Lymphoma | Lymp | C81–C85, C88, C96, C900, C902 |
| 19 | Metastatic Cancer | METS | C77–C80 |
| 20 | Solid Tumor without Metastasis | Tumor | C00–C26, C30–C34, C37–C41, C43, C45–C50, C51–C58, C60–C76, C97 |
| 21 | Rheumatoid Arthritis/Collagen | Rheum_A | L940–L941, L943, M05–M06, M08, M120, M123, M30–M35, M45, M461, M468–M469 |
| 22 | Coagulopathy | Coag | D65–D68, D691, D693–D696 |
| 23 | Obesity | Obesity | E66 |
| 24 | Weight Loss | WL | E40–E46, R634, R64 |
| 25 | Fluid and Electrolyte Disorders | Fluid | E222, E86, E87 |
| 26 | Blood Loss Anemia | BLA | D500 |
| 27 | Deficiency Anemia | DA | D508–D509, D51–D53 |
| 28 | Alcohol Abuse | Alcohol | F10, E52, G621, I426, K292, K700, K703, K709, T51, Z502, Z714, Z721 |
| 29 | Drug Abuse | Drug | F11–F16, F18–F19, Z715, Z722 |
| 30 | Psychoses | Psycho | F20, F22–F25, F28–F29, F302, F312, F315 |
| 31 | Depression | Dep | F204, F313–F315, F32–F33, F341, F412, F432 |

S. Table 2. Chief complaint categories

| **Category** | **Keywords** |
| --- | --- |
| **respiratory** | vascular, respiratory, breath, cough, wheeze, lung, nasal, throat, asthma, smoke inhalation, hypoxia, copd, shortness |
| **cardiac** | chest, heart, palpitations, cardiac, stroke, syncope, fatigue, heartbeat, heart rate |
| **mental_health** | panic, suicide, psychiatric, anxiety, depression, hallucination, mental, delirium, tremens, agitation, homeless, homicidal, withdrawal |
| **neurological** | head, dizziness, seizures, vomiting, weak, migraine, blurred, vision, cerebrospinal, parkinson, lethargy, numbness, droop, withdrawal |
| **gastrointestinal** | abdominal, vomit, diarrhea, nausea, stomach, cramp, gerd, stool, hemorrhoids, fecal, constipation, urine, vomiting, swallow, flank |
| **musculoskeletal** | back, shoulder, leg, arm, knee, joint, extreme, osteomyelitis, aches, hip, pain, wrist, ankle, neck, trauma |
| **genitourinary** | urine, vagina, penis, dysuria, kidney, rectal, rupture of membranes, contractions, ectopic pregnancy, pregnancy, pelvic, urinary, menstrual, vaginal |
| **dermatological** | rash, itch, hive, skin, swell, abscess, laceration, cellulitis, lip laceration, swelling, drainage, allergic |
| **overdose** | addiction, alcohol, intoxication, drug, overdose, withdrawal, substance use, poison |
| **pain/injury** | pain, injury, wound, bleeding, bleed, fall, battery, eye, burn, animal bite, laceration, dehiscence, ache |
| **ophthalmological** | eye, vision, blepharitis, blurred |
| **infectious_disease** | fever, flu, epidemic, jaundice, infection, body fluid exposure, chills, cold, sinusitis, covid |
| **other** | unspecified, abnormal, transplant, pregnancy, post-op, foreign body, hearing, dental, angioedema, throat |

S. Figure 1 – Feature importance scores for Random Forest

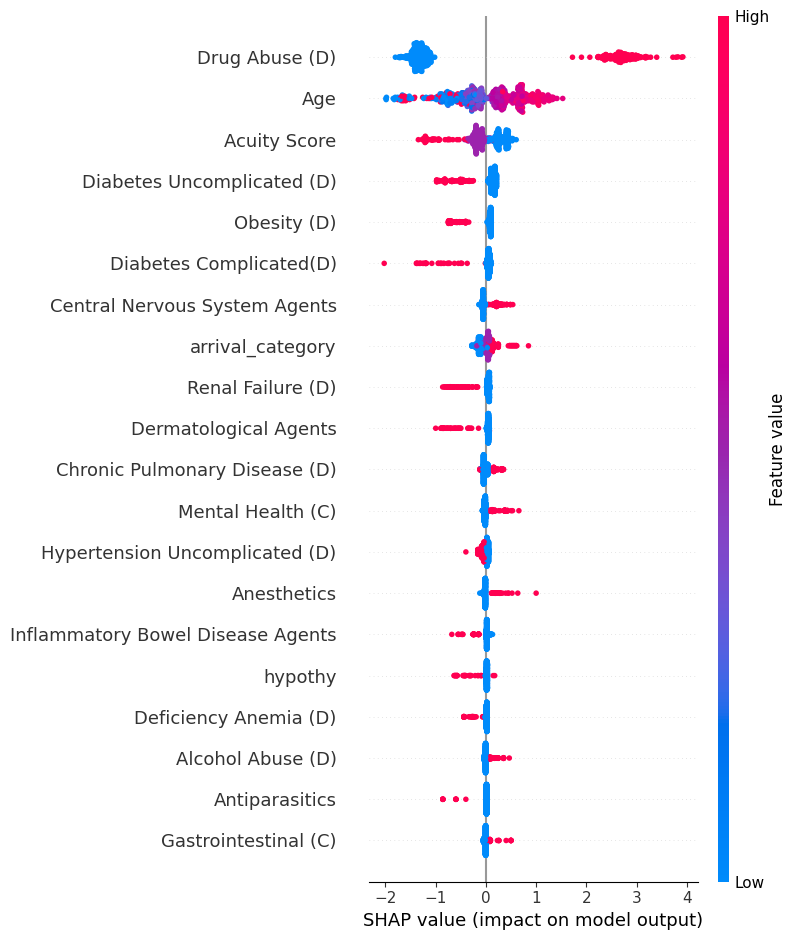

S. Figure 2 – SHAP-feature importance score for Neural Network

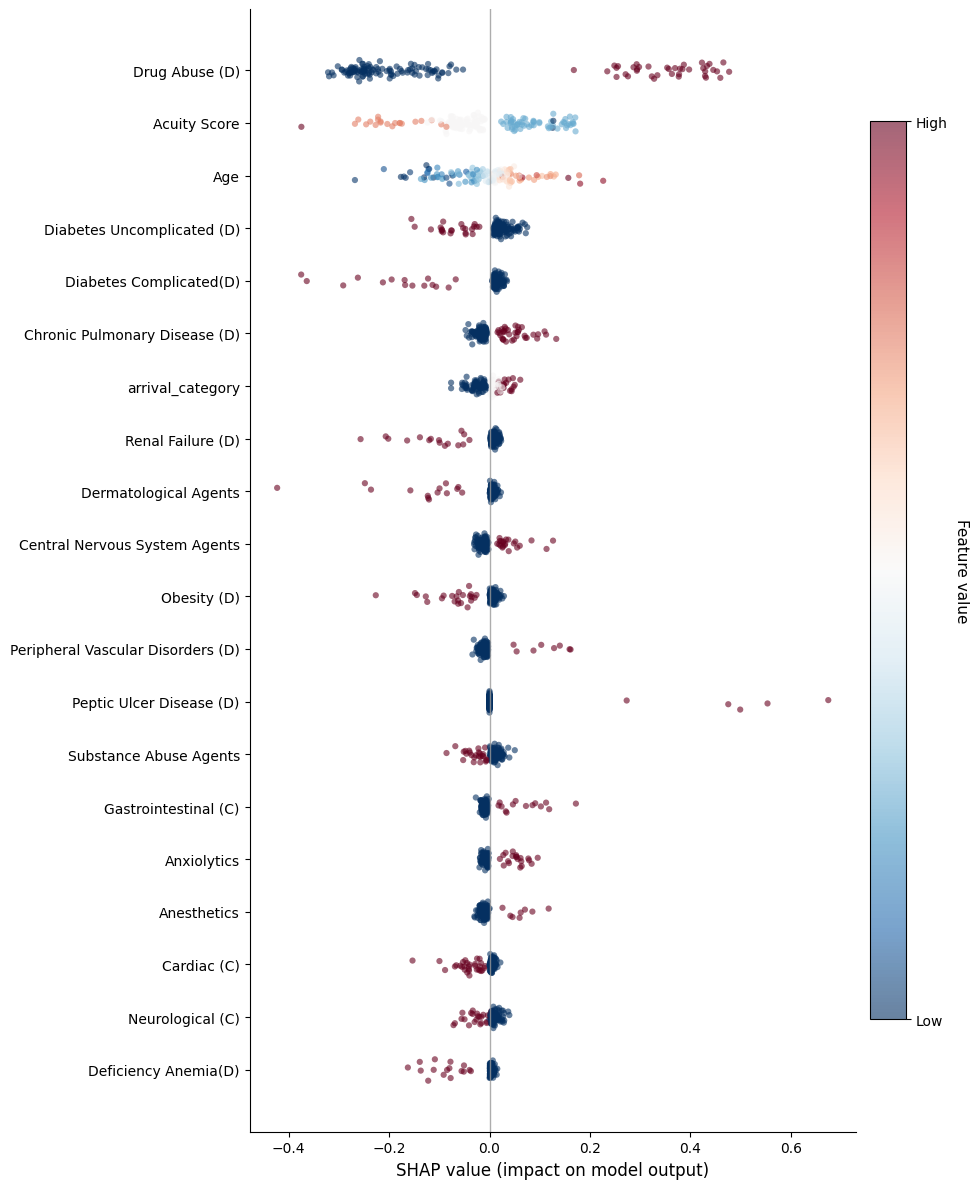

S. Table 3. Results with Mean value imputation for ED Acuity

| **Model** | **Accuracy** | **Precision** | **Recall** | **F-1 Score** | **AUROC** | **AUPRC** |
| --- | --- | --- | --- | --- | --- | --- |
| ICD-10-CM Baseline* | 0.87 | 0.96 | 0.73 | 0.83 | - | - |
| Logistic Regression | 0.74 | 0.75 | 0.73 | 0.74 | 0.86 | 0.86 |
| Support Vector Machine | 0.75 | 0.73 | 0.72 | 0.73 | - | - |
| Random Forest | 0.81 | 0.81 | 0.78 | 0.79 | 0.86 | 0.88 |
| Bagging | 0.76 | 0.75 | 0.71 | 0.73 | 0.80 | 0.81 |
| Gradient Boosting | 0.81 | 0.84 | 0.71 | 0.77 | 0.87 | 0.89 |
| K-Nearest Neighbors | 0.69 | 0.64 | 0.75 | 0.69 | 0.75 | 0.68 |
| Naive Bayes | 0.58 | 0.52 | 0.99 | 0.68 | 0.79 | 0.75 |
| AdaBoost | 0.78 | 0.72 | 0.74 | 0.75 | 0.84 | 0.86 |
| Decision Tree | 0.74 | 0.69 | 0.75 | 0.73 | 0.74 | 0.64 |
| Linear Discriminant Analysis | 0.76 | 0.73 | 0.70 | 0.72 | 0.82 | 0.80 |
| Quadratic Discriminant Analysis | 0.51 | 0.48 | 0.97 | 0.63 | 0.60 | 0.52 |
| Extra Trees | 0.79 | 0.73 | 0.71 | 0.72 | 0.82 | 0.83 |
| XGBoost | 0.76 | 0.78 | 0.67 | 0.72 | 0.85 | 0.81 |
| Feed Forward Neural Network | 0.78 | 0.79 | 0.76 | 0.77 | 0.79 | 0.75 |

S. Table 4. Results with K-nearest neighbors (k=3) value imputation for ED Acuity

| **Model** | **Accuracy** | **Precision** | **Recall** | **F-1 Score** | **AUROC** | **AUPRC** |
| --- | --- | --- | --- | --- | --- | --- |
| ICD-10-CM Baseline* | 0.87 | 0.96 | 0.73 | 0.83 | - | - |
| Logistic Regression | 0.74 | 0.75 | 0.73 | 0.73 | 0.86 | 0.86 |
| Support Vector Machine | 0.74 | 0.73 | 0.71 | 0.72 | - | - |
| Random Forest | 0.81 | 0.81 | 0.78 | 0.79 | 0.86 | 0.88 |
| Bagging | 0.76 | 0.75 | 0.71 | 0.73 | 0.80 | 0.81 |
| Gradient Boosting | 0.81 | 0.84 | 0.71 | 0.77 | 0.86 | 0.89 |
| K-Nearest Neighbors | 0.70 | 0.64 | 0.75 | 0.69 | 0.75 | 0.69 |
| Naive Bayes | 0.58 | 0.53 | 0.99 | 0.68 | 0.78 | 0.74 |
| AdaBoost | 0.78 | 0.72 | 0.74 | 0.75 | 0.84 | 0.86 |
| Decision Tree | 0.74 | 0.69 | 0.75 | 0.73 | 0.74 | 0.64 |
| Linear Discriminant Analysis | 0.77 | 0.73 | 0.71 | 0.72 | 0.82 | 0.80 |
| Quadratic Discriminant Analysis | 0.51 | 0.48 | 0.97 | 0.63 | 0.60 | 0.52 |
| Extra Trees | 0.79 | 0.73 | 0.71 | 0.72 | 0.82 | 0.83 |
| XGBoost | 0.76 | 0.78 | 0.67 | 0.71 | 0.85 | 0.81 |
| Feed Forward Neural Network | 0.78 | 0.79 | 0.76 | 0.77 | 0.79 | 0.75 |

S. Table 5. Results for Random forest classification model upon varying decision boundary of the classifier.

| Decision Boundary | Accuracy | Precision | Recall | F1 Score | AUROC | AUPRC |
| --- | --- | --- | --- | --- | --- | --- |
| 0.05 | 0.46 | 0.46 | 1 | 0.63 | 0.87 | 0.88 |
| 0.1 | 0.48 | 0.47 | 1 | 0.64 | 0.87 | 0.88 |
| 0.15 | 0.49 | 0.47 | 0.99 | 0.64 | 0.87 | 0.88 |
| 0.2 | 0.53 | 0.49 | 0.97 | 0.65 | 0.87 | 0.88 |
| 0.25 | 0.6 | 0.54 | 0.94 | 0.68 | 0.87 | 0.88 |
| 0.3 | 0.66 | 0.58 | 0.93 | 0.72 | 0.87 | 0.88 |
| 0.35 | 0.71 | 0.62 | 0.91 | 0.74 | 0.87 | 0.88 |
| 0.4 | 0.78 | 0.71 | 0.88 | 0.79 | 0.87 | 0.88 |
| 0.45 | 0.79 | 0.76 | 0.78 | 0.77 | 0.87 | 0.88 |
| 0.5 | 0.79 | 0.8 | 0.71 | 0.75 | 0.87 | 0.88 |
| 0.55 | 0.8 | 0.85 | 0.68 | 0.76 | 0.87 | 0.88 |
| 0.6 | 0.79 | 0.88 | 0.64 | 0.74 | 0.87 | 0.88 |
| 0.65 | 0.78 | 0.89 | 0.59 | 0.71 | 0.87 | 0.88 |
| 0.7 | 0.77 | 0.93 | 0.55 | 0.69 | 0.87 | 0.88 |
| 0.75 | 0.74 | 0.94 | 0.46 | 0.62 | 0.87 | 0.88 |
| 0.8 | 0.71 | 1 | 0.38 | 0.55 | 0.87 | 0.88 |
| 0.85 | 0.69 | 1 | 0.32 | 0.48 | 0.87 | 0.88 |
| 0.9 | 0.63 | 1 | 0.2 | 0.34 | 0.87 | 0.88 |
| 0.95 | 0.57 | 1 | 0.07 | 0.14 | 0.87 | 0.88 |

S. Figure 3. Calibration Curve for Neural Network.

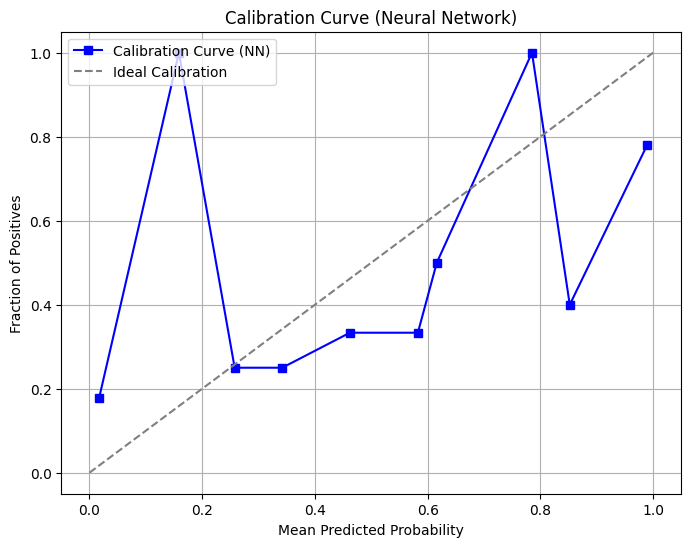

S. Figure 4. Calibration Curve for Random Forest.

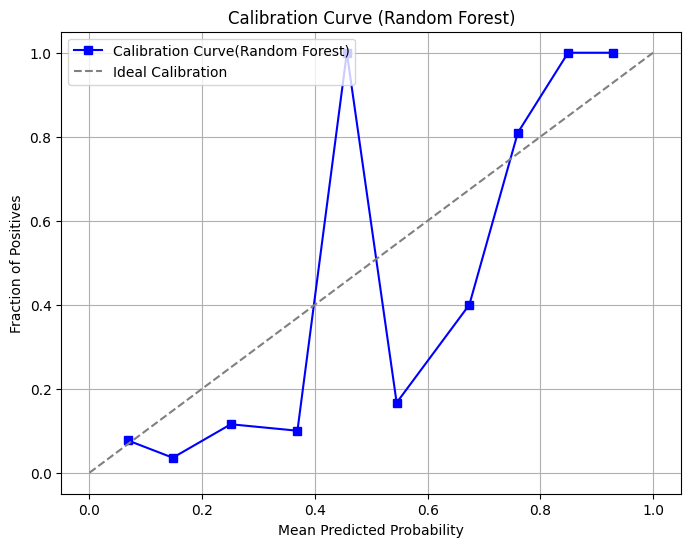

S. Figure 5. Calibration Curve for Gradient Boosting.

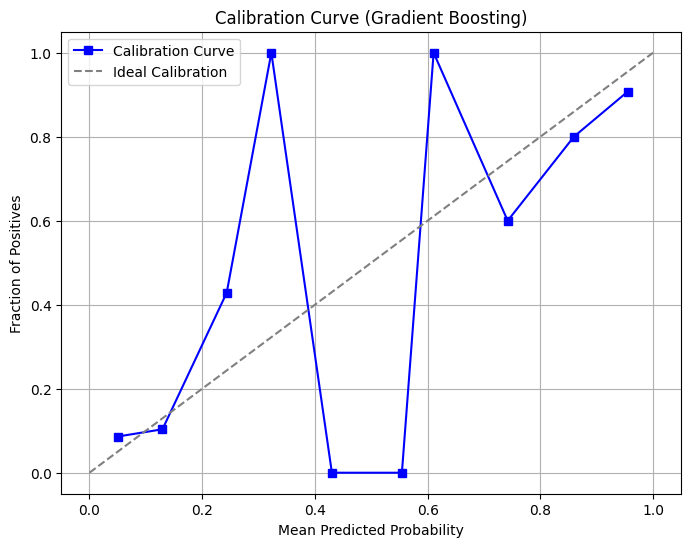

S. Figure 6. Calibration Curve for K-nn.

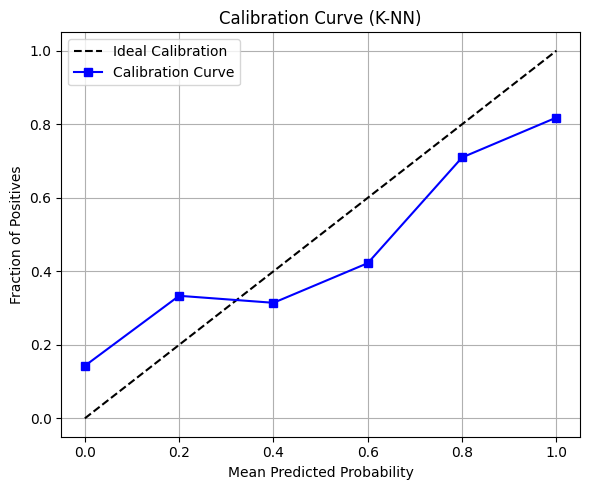

S. Figure 7. Calibration Curve for Logistic Regression

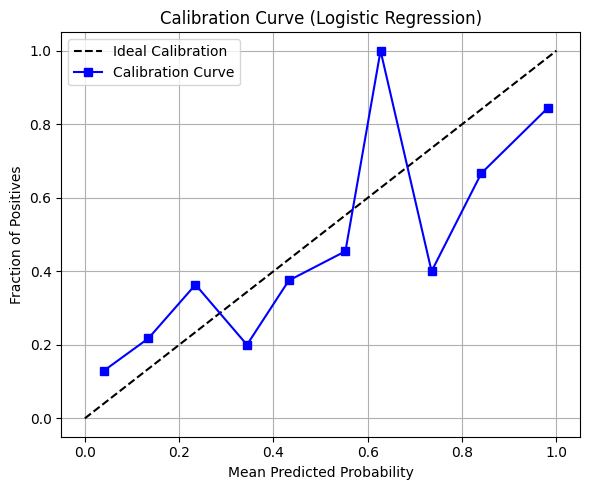

S. Figure 8. Calibration Curve for Bagging

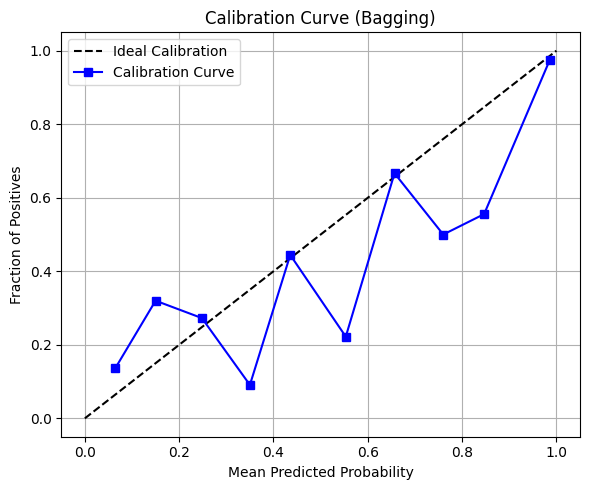

S. Figure 9. Calibration Curve for XGBOOST

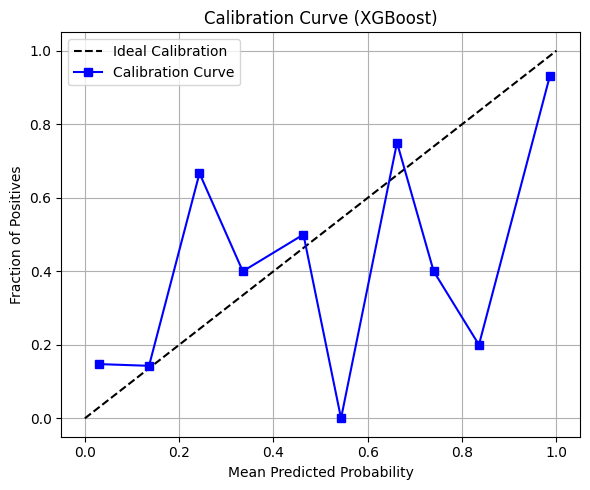

S. Figure 10. Calibration Curve for Naive Bayes
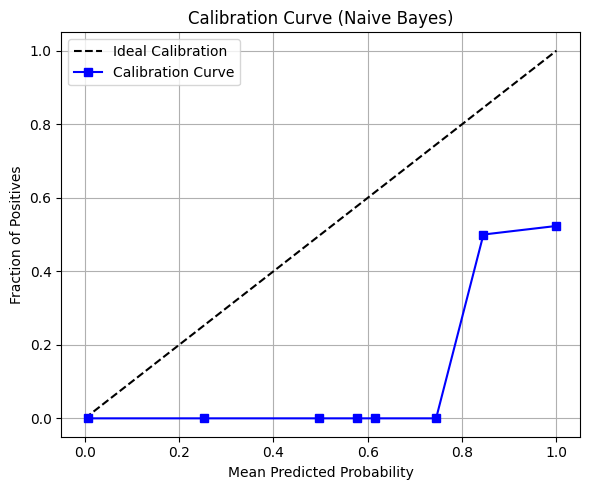

S. Figure 11. Calibration Curve for Quadratic Discriminant Analysis

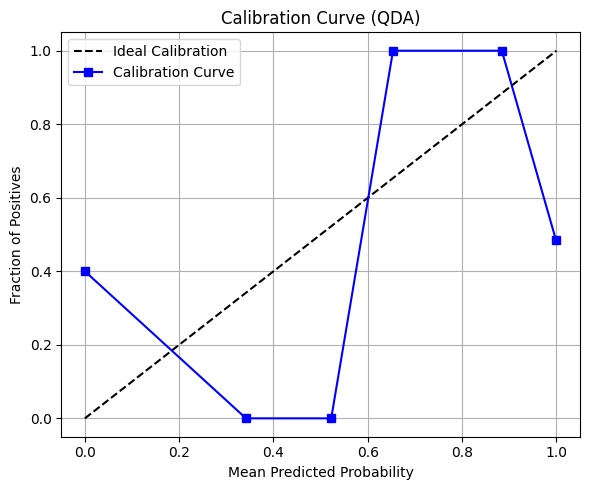

S. Figure 12. Calibration Curve for LDA

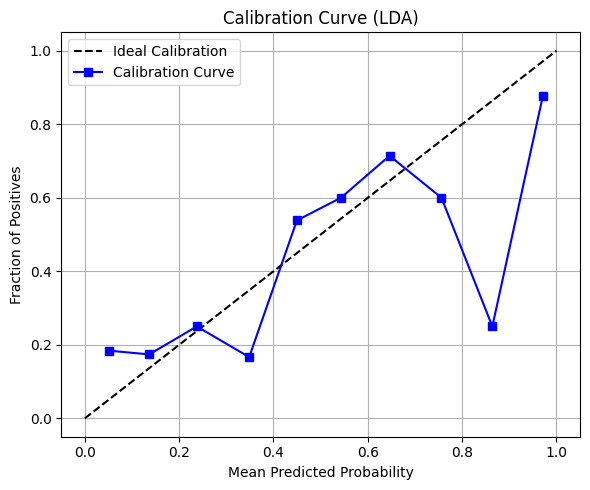

S. Figure 13. Calibration Curve for Extra Trees

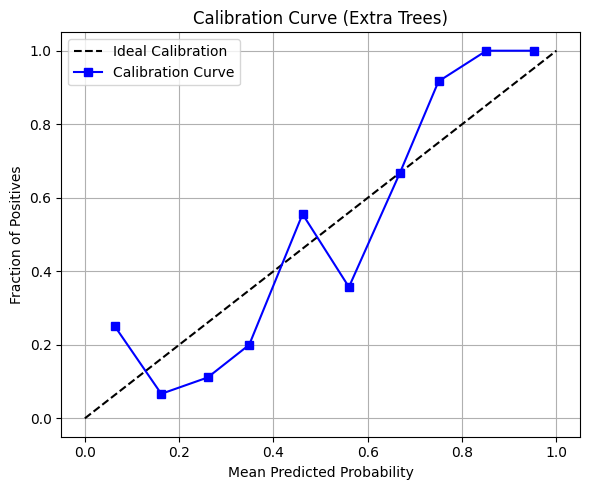

S. Figure 14. Calibration Curve for SVM Classifier.

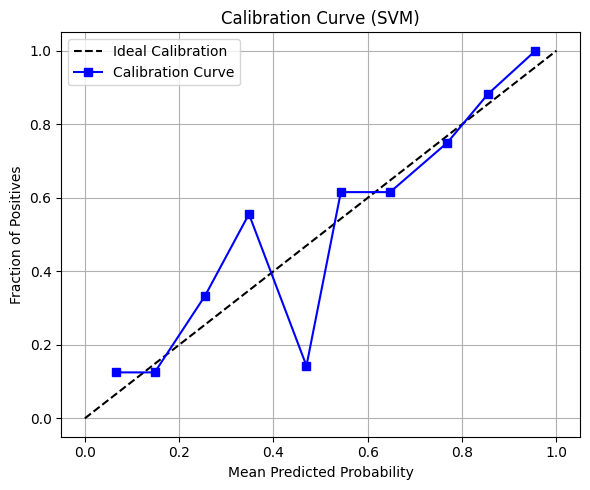
.

S. Table 5. Sklearn parameters for the different functions.

| **Model Name** | **Class Used** | **Key Parameters** |
| --- | --- | --- |
| Logistic Regression | LogisticRegression() | Default |
| Support Vector Machine | SVC() | Default |
| Random Forest | RandomForestClassifier() | Default |
| Bagging | BaggingClassifier() | estimator=base_estimator, n_estimators=15, random_state=42 |
| Gradient Boosting | GradientBoostingClassifier() | n_estimators=15, random_state=20 |
| K-Nearest Neighbors | KNeighborsClassifier() | Default |
| Naive Bayes | GaussianNB() | Default |
| AdaBoost | AdaBoostClassifier() | Default |
| Decision Tree | DecisionTreeClassifier() | Default |
| Linear Discriminant Analysis | LinearDiscriminantAnalysis() | Default |
| Quadratic Discriminant Analysis | QuadraticDiscriminantAnalysis() | Default |
| Extra Trees | ExtraTreesClassifier() | n_estimators=100, random_state=42 |
| XGBoost | XGBClassifier() | n_estimators=100, random_state=42 |
| Neural Network (PyTorch) | ImprovedNeuralNet | See Below |

S. Table 6. Neural Network parameter

| **Layer Type** | **Details** |
| --- | --- |
| Input Layer | Linear(input_size, 128) |
| BatchNorm 1 | BatchNorm1d(128) |
| Activation | ReLU() |
| Dropout | Dropout(0.3) |
| Hidden Layer 2 | Linear(128, 64) |
| BatchNorm 2 | BatchNorm1d(64) |
| Activation | ReLU() |
| Dropout | Dropout(0.3) |
| Hidden Layer 3 | Linear(64, 32) |
| Activation | ReLU() |
| Output Layer | Linear(32, 2) (2 classes) |
